# Smart Testing with Vaccination: A Bandit Algorithm for Active Sampling for Managing COVID-19

**DOI:** 10.1101/2021.05.01.21256469

**Authors:** Yingfei Wang, Inbal Yahav, Balaji Padmanabhan

**Affiliations:** Foster School of Business, University of Washington; Coller School of Management, Tel Aviv University; Muma College of Business, University of South Florida

**Keywords:** COVID-19, active sampling, multi-armed bandit, agent-based models, smart testing

## Abstract

This paper presents methods to choose individuals to test for infection during a pandemic such as COVID-19, characterized by high contagion and presence of asymptomatic carriers. The smart-testing ideas presented here are motivated by active learning and multi-armed bandit techniques in machine learning. Our active sampling method works in conjunction with vaccination and quarantine policies and is adaptive to changes in real-time data. Using a data-driven agent-based model simulating New York City we show that the algorithm samples individuals to test in a manner that rapidly traces infected individuals. The results show that smart-testing is effective in significantly reducing infection and death rates as compared to current policies, with or without vaccination.

## 1 Introduction

We present a method to actively sample individuals to test as a way to mitigate the spread of pandemics such as COVID-19. Sampling algorithms are common in machine learning to acquire labels for classification (“active learning”, Cohn et al. (1996)) and in bandit algorithms, (Bubeck and Cesa-Bianchi 2012) to explore complex search spaces through exploration and exploitation. In public health, there are two reasons why the literature has considered sampling. The first is to estimate the actual spread of a disease, such as HIV using sentinel-based strategies (e.g., Magnani et al. 2005). The second is to mitigate the spread of disease by proactively identifying individuals who need to be quarantined or tested. One method to do this is contact-tracing (CT) (Eames and Keeling 2003), which is the main mechanism used worldwide during COVID-19 (Hellewell et al. 2020).

The SARS-Cov-2 pathogen and its transmission have the following unique characteristics, though, that make none of these sampling methodologies sufficient by itself.

- The possibility of asymptomatic spread rules out pure sentinel-based strategies that would test those who report symptoms. For the same reason, contact tracing alone would be insufficient. This is because contact-tracing pursues only contacts of those known to have tested positive. Asymptomatic carriers may have also been transmitting the disease to their contacts, but those will not be actively sought by this strategy.
- The pathogen has also been shown to survive on surfaces or in air in specific locations for extended periods of time. Hence, location-based sampling may also be useful.
- Information about how covariates might affect contracting the disease is still uncertain. In this environment, covariate-based sampling strategies alone might be insufficient. Sampling strategies that target specific *individuals* to test is therefore critical.
- The highly infectious nature of the pathogen and the extent of worldwide spread have placed constraints on testing capacities, requiring the prioritizing of tests.

We address all four aspects noted above. This paper presents a dynamic active sampling strategy based on the multi-armed bandit algorithm that can optimally combine the different sampling ideas to generate real-time lists of whom to sample. Bandit algorithms are particularly good at combining exploration with exploitation and have seen success in many scenarios where this combination is important. This is the case with SARS-Cov-2, where exploration (random sampling) has to be effectively combined with exploitation (contact-tracing, location-based sampling) in a dynamic manner based on data. One of the key aspects of our work is that we identify specific individuals to test/sample, as opposed to criteria or covariate-based sampling strategies.

Our algorithm operates in a high uncertainty network environment; The contacts between individuals are initially only partially observable and are revealed gradually based on individual sampling. The bandit algorithm therefore first uses the Thompson sampling strategy to trade off between expansion in unobserved nodes to identify possible new hot spots, vs. proliferate in the observed portion to utilize the knowledge learnt for testing. If proliferation is chosen, an inner level upper confidence bounding (UCB) policy is designed to balance between the individuals having higher probability of infection vs. those having greater information uncertainty.

Recent COVID-19 studies have used agent-based models (ABMs) with standard S/I/R-based diffusion models to study various policies (e.g., Keskinocak et al. 2020). An ABM of the COVID-19 spread in New York was presented in Hoertel et al. (2020). This model serves as a basis for the ABM that we developed in this work, and reported in Appendices A-B.

There is a similar question of whom to vaccinate (Chen et al. 2020). Our framework is general and allows for various vaccination policies to be incorporated into the background dynamics in order to test the effectiveness of combinational strategies such as smart testing with vaccination. Given the importance of this today, our experimental results focus on combining smart-testing and vaccination policies.

We make the following important contributions. First, we present a novel active sampling algorithm to effectively manage pandemics such as COVID-19. We develop a general multi-armed bandit framework for this problem that (a) can leverage information of different types such as individuals and locations, (b) can handle uncertainty in both the underlying disease dynamic as well as information and (c) is built to sample based on individuals and not covariates. Second, we make important contributions to the literature on multi-armed bandits as well. Beyond classic bandit problems, there exists limited literature on active search on graph, with the objective of finding as many target nodes as possible with some given property. Most of the existing work assumes that the complete network structure and underlying process is known beforehand. There is very little work on partially observed and dynamically changing networks and underlying (disease) dynamics. We model the partially observed scenarios within this framework and show how the network can be strategically expanded over time to support the active sampling strategy. Third, we show that smart testing is still important even when vaccination is available, assuming that some portion of the individuals cannot or will not get the vaccine in a timely manner. Further, pandemics such as COVID-19 are likely going to recur in the future, with no guarantees that vaccines and/or massive tests will be immediately available. In this paper, we view vaccines as complementary to testing, and, as we show, active testing strategies remain important even with vaccinations as a combinational strategy to eradicate pandemics.

## 2 Problem Formulation

Taking a policy-maker’s perspective, our setting has three components. The *data setting* represents knowledge about individuals and their spatio-temporal behavior. The *process setting* represents knowledge about the underlying disease spread. The *policy setting* represents what policy makers are assumed to have access to, and accordingly what the sampling algorithm will use. These are described below.

Data setting:

- a population of individuals *P*,
- a set of locations *L* in the city,
- spatial-temporal data about individuals in *P* over *L*.

Process setting:

- initial actual disease state (e.g., S/I/R) of each individual in *p* ∈ *P*,
- a spatio-temporal model of contagion that represents how the disease spreads in the population,

The policy-makers’ setting involves two components, the information setting and the policy setting. The information setting includes:

- complete list of individuals *P*,
- complete list of locations *L*,
- initial disease state of each individual in *P* (this can be incomplete),
- the contagion process is partially known - the policy maker (algorithm) is assumed to be aware of only the high-level factors affecting disease diffusion, such as contacts with exposed individuals or the role of locations where infected individuals have been.

The policy setting includes:

- a quarantine policy that determines when and how individuals will isolate in the population,
- a vaccination policy that determines which and when individuals will receive vaccine,
- an objective function *O*(*T*). This paper focuses on sampling strategies for bridging the gap between the actual infected population and the known infected individuals, as a step towards controlling disease spread.
- constraints on number of tests per day (*D*_*max*_).
- Sampling an individual to test reveals the following:
  – disease state (infected or not) of the individual being tested.
  – spatio-temporal data about the individual, if infected. In some cases (e.g. access to full GPS data) this will be complete information on the places visited and the actual contact network. If this is by self-reporting (as is common in most places) then this information will be incomplete.

Given this, the policy maker seeks to sample individuals for testing in order to optimize the objective.

## 3 Active Sampling Framework and Algorithm

The Multi-Armed Bandit (MAB) is a generic framework to address the problem of decision making under uncertainty. In this setting, the learner must choose from among a variety of actions and only observes partial feedback from the environment, without prior knowledge of which action is the best. In the classic stochastic *K*-armed bandit problem, at each time step *t*, the learner selects a single action/arm *x*_*t*_ among a set of *K* actions and observes some payoff *r*_*t*_(*x*_*t*_). The reward of each arm is assumed to be drawn stochastically from some unknown probability distribution. The goal of the learner is maximize the cumulative payoff obtained in a sequence of *n* allocations over time, or equivalently minimize the *regret* (Bubeck and Cesa-Bianchi 2012), which is defined as the difference between the cumulative reward obtained by always playing the optimal arm and the cumulative reward achieved by the learning policy,

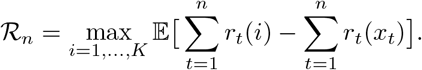

The fundamental exploration/exploitation dilemma is to: (1) gain as many rewards as possible in the current round, but also (2) have a high probability of correctly identifying the better arm. Beyond classic bandit problems, there exists literature on active search, with the objective of finding as many target nodes as possible with some given property. Most of the existing work assumes that the complete network structure is known before hand (e.g., Ma et al. 2015), with the exception of a few recent papers that consider partially observed networks (e.g., Singla et al. 2015, Soundarajan et al. 2017, Madhawa and Murata 2019). However these approaches do not consider the node sampling scenario needed for the testing problem considered in this paper.

In the MAB context of active search on graphs, the question our research addresses can be posed, more generally, as: *given a partially observed network with no information about how it was observed, and a budget to query the partially observed network nodes, can we learn to sequentially ask optimal queries?* This is a novel extension to the MAB literature. For this problem we offer an important methodological contribution that uses network embedding ideas with two-levels of exploration/ exploitation tradeoffs. We also offer important modeling contributions in order to be able to cast the smart testing problem in a MAB framework as we also show below.

### 3.1 The Sampling Framework

We model contact networks as an undirected, time dependent, network 𝒢_*t*_ =*< V, E, D >*, where *V* is the node representation of the individuals *P*. The network is evolving over time with new contacts reported to the policy maker. For example, if the policy maker aims to make daily decisions, 𝒢_*t*_ represents the daily snapshot of the network on day *t*. An edge *e*_*ij*_ ∈ *E* represents a direct contact between individuals *v*_*i*_ and *v*_*j*_. At any time point, every node *v* in our graph 𝒢_*t*_ has one disease state in *D*. We use *y*_*v*_ to represent whether the individual is tested positive. The actual disease state can never be fully observed (e.g. the difference between susceptible, exposed and recovered cannot be identified).

The graph 𝒢_*t*_ is not fully observable and only partial information 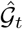 is available to the decision maker. When a node tests “positive”, its *known* contacts will be revealed at once, with the understanding that there might be other contacts that are latent and not readily observed.

Mathematically, this problem of active sampling can be formalized as a stochastic combinatorial optimization problem with *< V*, U, *f >*, where *V* is the set of nodes to be sampled from, U = {*U* ⊂ *V* : |*U* | ≤ *D*_*max*_} is a family of subsets of *V* with up to *D*_*max*_ (daily testing capacity) nodes, and *f* a function that maps the node to [0,1] as the probability of infection. The objective function for all the nodes in a set *U* ⊂ *V* is defined by *F* (*U, f*). For example, if the objective is to find as many infected nodes as possible in each day, *F* (*U, f*) can be defined as:

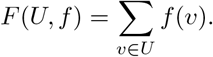

The goal of the sampling policy is to adaptively select a sequence of subsets *U*_*t*_ to test, in a way that maximizes the cumulative rewards of *F* (*U*_*t*_, *f*) over time, recognizing that we observe, on testing, the realized reward *f* (*v*) of each node *v* ∈ *U*_*t*_ immediately, i.e. whether the tested individual is infected or not (*y*_*v*_).

An important observation is that immediate contacts alone (as done in previous work (e.g., Singla et al. 2015, Soundarajan et al. 2017, Madhawa and Murata 2019)) may be insufficient to consider for COVID-19 testing and we need a richer representation of similarity between individuals. We seek to model *similarity* such that if one agent is infected, then a *similar* node is also possibly infected. This can help identify potential regions at high risk of outbreaks given previous outbreak locations, the mobility of agents, and their network structure. Local structure features, like degree, number of triads, and centrality, have been used in previous network analysis on finding structurally similar nodes (Henderson et al. 2012). Although these features can help infer roles of each node, such as super spreaders, or periphery nodes, they do not capture the information about the neighborhood similarity, social relations, and community membership. For example, if two agents are living in the same household, COVID-19 can easily spread through contact transmission or droplet transmission. Due to large amount of presymptomatic and asymptomatic disease transmissions, even if there is no direct contact link between two agents, if they constantly went to the same supermarket, or live in nearby neighborhoods, the agents can end up infecting each other. This complex interplay of social relationships and locations has resulted in several infections from the same gathering (super-spreaders, hotspots) and brought up the importance of even seemingly minor occurrences such as how even a few agents with the same travel history may have caused unknown breakouts along the way.

Our goal is to quantify these above mentioned social relations by a continuous latent representation of nodes, which can then be exploited to guide the sampling policy to more effectively allocate testing kits of limited capacity. A pioneering method, DeepWalk (Perozzi et al. 2014), uses language modeling approaches to learn latent node embedding in the following two steps (please refer to Appendix C for the technical details of this approach). First it traverses the network with random walks to infer local structures by neighborhood relations, and then uses a SkipGram model (Mikolov et al. 2013) to learn node embedding based on the produced samples. In our context, by translating the nodes in the network into a continuous space this way, the node embedding thus generated provides a method to efficiently sample individuals to test based on how “close” they are to others.

### 3.2 The Sampling Algorithm

The sampling algorithm faces two major challenges: (1) the policy maker (through contact-tracing from positive tests) observes only a portion of the network at any time *t* (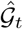, as illustrated in Fig. 1(a)), and (2) sampling individuals, as opposed to group sampling, entails an exponential number of possible arms to sample with the cardinality constraint *D*_*max*_ (daily testing capacity).

**Figure 1:**
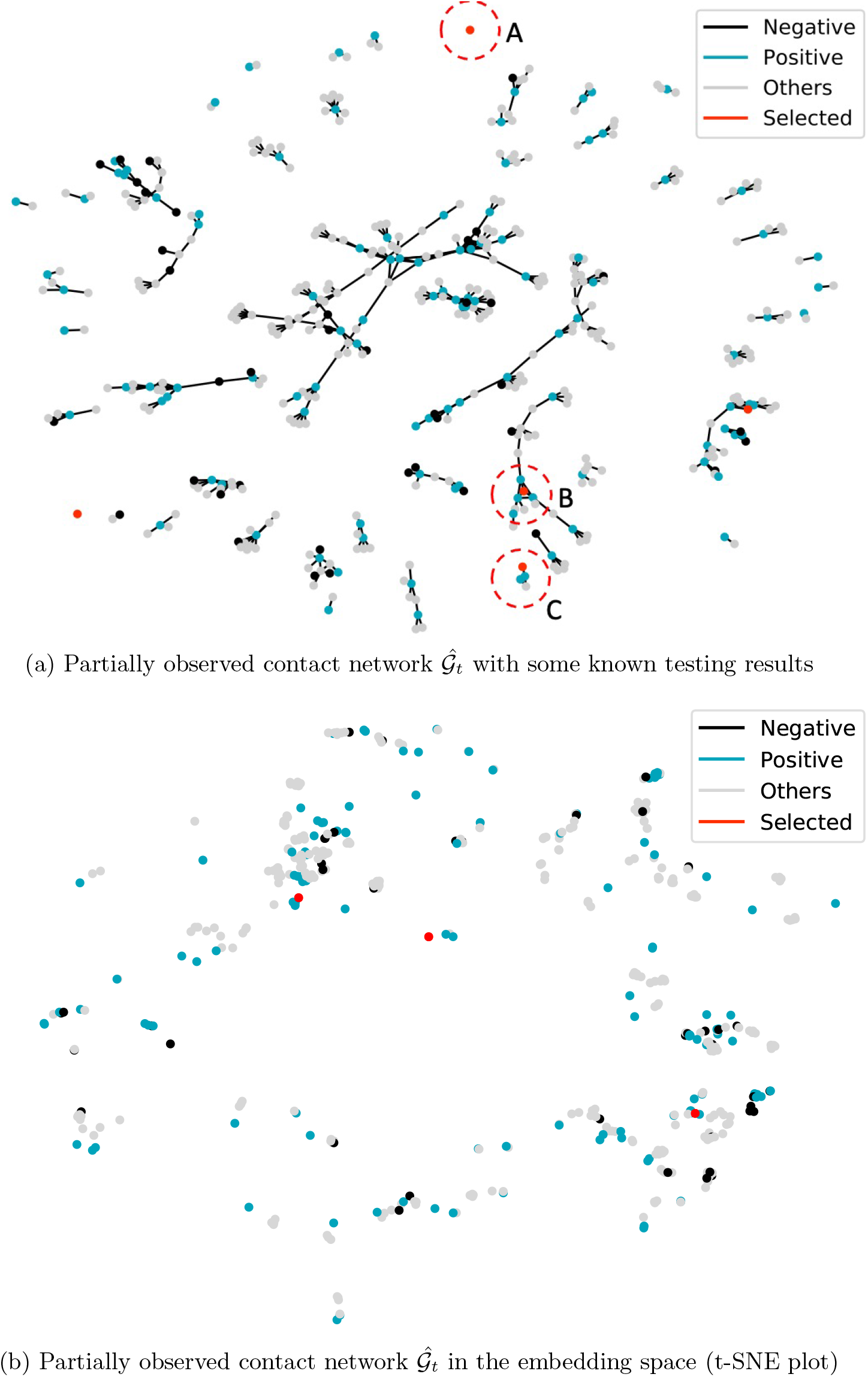
An illustrative example of the partially observed contact network, and the two levels of exploration/exploitation trade-off. Known testing results are reported, together with the individuals selected by the proposed sampling policy.

There are, in turn, three sources of uncertainty. First, whether the observed contacts are infected is not known, and needs to be learnt as we sample individuals and observe their test results. Second, since the contact network is only partially observed, other possible paths of disease transmissions are not known to the policy maker. Third, the vast majority of the population is not reported in the current contact network 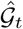, and it is worthwhile to observe a broader area of the underlying network to detect and control possible outbreaks in unreported areas.

Hence the proposed sampling policy concerns two levels of exploration/exploitation tradeoff.

#### Outer Level: expansion vs. proliferation

In regard to the third source of uncertainty, the outer level is modeled as a two-armed bandit, balancing between the two choices of expansion outside the current known contact network, and/ or proliferation within the current network. For example, in Fig. 1(a), individual *A* is not observed in the current contact network, and is selected by the active sampling policy based on the choice of expansion.

We choose to use the Thompson sampling policy with Beta-Bernoulli distribution (Thompson 1933, Chapelle and Li 2011). The expected reward *θ* of the two choices are modeled with a Beta distribution. After one choice is chosen, the realized reward (whether an individual is tested positive) is sampled from a Bernoulli distribution. We repeatedly sample *D*_*max*_ realizations and choose between the two choices of expansion/proliferation that has the highest sampled value. If expansion is chosen, our policy samples a node *v* that was not in 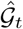 uniformly at random. If proliferation is chosen, our policy uses the inner level exploration/ exploitation algorithm to choose individuals *v* to sample. We use 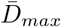 to represent the total number of tests that are allocated to proliferation. After the test result *y*_*v*_ for each chosen individual is revealed, the posterior Beta distribution will be updated, according to whether node *v* is from expansion or proliferation.

#### Inner Level: individual selection within the current observed network (combinatorial *k*NN-UCB)

Within the current observed contact network, the first two sources of uncertainty give rise to a trade-off between choosing individuals that are *seemingly* more prone to be infected, and/or learning the network to get more contact information to detect more infected in the future. As illustrated in Fig. 1(a), given the current observed network, the probability of individual *B* is infected is very high since three of his/her direct contact was tested positive, together with other confirmed individuals nearby. In comparison, the neighboring information of individual *C* is very limited, rendering in higher uncertainty. It might be possible that individual *C* has a higher chance of being infected and we need to learn his/her surroundings.

The goal of the inner level bandit algorithm is to effectively allocate 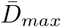 lab tests to individuals to alleviate the disease spread. Specifically, it adaptively selects a sequence of subsets *U*_*t*_ ⊂ *V* to test and maximize the cumulative rewards of *F* (*U*_*t*_, *f*) over time. We also observe the realized reward *f* (*v*) of each node *v* ∈ *U*_*t*_, i.e. whether the tested individual is infected or not.

Thus, to balance the exploitation/exploration within the current observed network, on each day, we first use network embedding *x*_*v*_ ∈*χ* (*χ* ⊂ℝ^*d*^) from Section 3.1 to find the node representation. The node embedding encodes the network structure, i.e. neighbors and connection information, and quantifies the similarity between two nodes in the sense that if one is infected then the other is highly likely to be infected from potential contact chains. Fig. 1(b) is an illustration of the observed nodes in the embedding space, which translates the network structure to continuous space for direct sampling from the entire population. In our case, each individual is represented by a lower dimensional embedding (“lying in a manifold”). In this case, *k*NN regression can have provable bounds (Jiang 2019).

##### Definition 1

(*k*NN). *Let the kNN radius of x* ∈*χ* ⊂ℝ^*d*^ *be ρ*_*k*_(*x*) := inf{*ρ* : |*B*(*x, ρ*) ∩*χ* | ≥ *k*}, *where B*(*x, ρ*) *denotes the open metric ball of radius ρ >* 0, *centered at x. Let* 𝒩_*k*_(*x*) *denote the k-neighborhood of x, such that* 𝒩_*k*_(*x*) := *B*(*x, ρ*_*k*_(*x*))∩ *χ*. *Then, for all x* ∈*χ, the kNN regressor is defined with respected to the training data* (*x*_1_, *y*_1_), …, (*x*_*n*_, *y*_*n*_) *as*

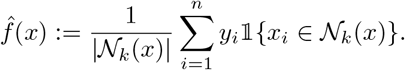

In order to construct a valid upper confidence bound, we follow the strong uniform consistency theorem for *k*NN regression.

##### Theorem 1

(Uniform Consistency (Jiang 2019)). *Let δ >* 0. *There exists N*_0_ *and universal constant C such that for n* ≥ *N*_0_ *and k* = *n*^2*/*(2+*d*)^, *then*

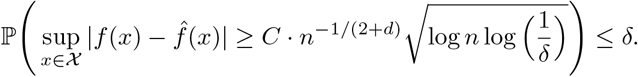

As illustrated in Fig. 1, on each day *t*, some of the testing results (negative/positive) are known to the policy maker, either by self-reporting, or by the active sampling policy. It is also worth noting that the network embeddings for two unconnected nodes are not comparable. Thus, when constructing the *k*NN prediction for each node, we need to first find the reported (either positive or negative) individuals that are connected to the focal node. At the same time, based on the uniform consistency theorem, the size of the neighborhood (the value of *k*) needs to be node-dependent.

We now describe our Combinatorial *k*NN-UCB as follows. On day *t*, for each node *v*_*i*_ in the current contact network, we use *N*_*t*_(*i*) to denote the number of known individuals (either tested positive or negative) that are connected to the focal node *v*_*i*_. The neighborhood size of the focal node is chosen as *k*_*t*_(*i*) = (*N*_*t*_(*i*))^2*/*(2+*d*)^. We use 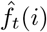 to denote the *k*NN estimate of the probability of infection of the focal node *v*_*i*_. At each time step, if only one individual will be sampled, based on the principle of *optimism in the face of uncertainty* (Auer et al. 2002), *the Upper Confidence Bound (UCB) policies define the largest plausible estimate* of each individual node as

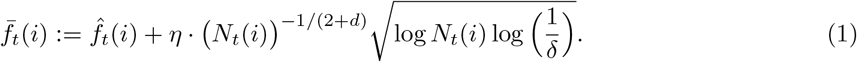

However, the high probability bound in Theorem 1 only holds for larger *n > N*_0_. To ensure that the upper confidence bound decreases with respect to the size *N*_*t*_(*i*) of the neighborhood, we modified the UCB for any 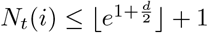 to be 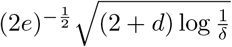, based on Theorem 2 (with proof provided in Appendix D). Then the algorithm chooses the arm that maximizes the above quantity.

##### Theorem 2.

*For* ∀*c >* 0, *α >* 0, 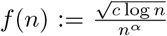 *is maximized at n*_0_ = *e*^1*/*2*α*^ *and is strictly decreasing for* ∀*n > n*_0_.

However, as mentioned above, in our specific COVID-19 active testing scenario, allocating 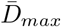 lab tests to individuals is a case of combinatorial optimization. To deal with this challenge, we allow the policy maker to use any exact/ approximation/ randomized algorithm, termed as ORACLE, to find solutions for the corresponding offline combinatorial optimization problem (if all the rewards *f* (*v*) were known beforehand to the policy maker),

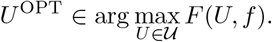

We denote the solution as 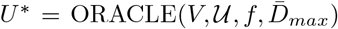 Notably *f* (*v*) *′s* are partially known to the policy maker. We thus propose the algorithm Combinatorial *k*NN-UCB which makes use of a greedy 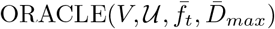 to provide (approximate) solutions for the offline optimization problem, with ties breaking randomly.

The confidence level *δ* should be small enough to ensure optimism with high probability, but not so small that the larger upper confidence bounds will result in excessive exploration of sub-optimal arms. We thus choose 1*/δ* = 1 + *t* log^2^(*t*), where *δ* is decreasing to zero slightly faster than 1*/t* to ensure sub-linear regret, and which is shown to improve the regret and empirical performance (Lattimore and Szepesvári 2020). The pseudocode is provided in Algorithm 2 in Appendix D.

## 4 Empirical Results

We compare three policies: Contact Tracing (CT), Random testing with CT and Active Sampling with CT as presented here. Among these options, CT is the current baseline in the US since random or active testing are currently not implemented. In the three scenarios we assume that symptomatic individuals are self quarantined, and contacts of infected individuals with a positive test quarantine through standard contact tracing processes.

We implemented active testing strategies under different vaccination scenarios and show that a combinational strategy is critical. We use a random vaccination policy that runs parallel to active testing and consider different extents of vaccination that eventually (after 100 days) reach to either *κ* = 25% or *κ* = 50% of the population. We chose these to closely reflect current numbers in the US (4 months after the start of vaccinations the US had fully vaccinated 25% of the population, and the annual percentage of those vaccinated for flu each year is approximately 50%). All of these can of course be varied as needed to reflect realities in different countries. We further vary the compliance level of individuals to self-isolate and quarantine in our empirical study (*γ* ∈ {10%, 50%}). Unfortunately there is no scientific consensus yet on the percentage of the US population who comply. We discuss the implications of this further below.

Consistent with recent daily tests in the US as reported in The Covid Tracking Project, we set daily testing capacity to be 0.5% of the population. We simulate each policy for 50 runs, under each of four scenarios, i.e. low/high compliance and low/high vaccination rate. We also simulate the corresponding scenarios without any vaccination in place. We simulate the disease spread for 350 days until the spread terminates. To account for the fact that testing tool-kits and vaccination are not developed at the beginning of the pandemic, we start the testing and vaccination on day 30. The running time is around 126 hours for 50 runs, under AWS EC2 r4.4xlarge instance, with 122 GiB memory and 4x 2.3GHz Intel Xeon E5-2686 v4 Processors.

We report our study results in Figures 2 and 3 and in Tables 1 and 2. Fig. 2 reports the simulation results under high compliance level and low vaccination rate, averaged over 50 runs. The left column depicts, 1) the percentage of the allocated tests returned with a positive result, 2) daily confirmed new symptomatic patients, 3) daily new infected, and 4) cumulative infected within the entire population. Due to the fact that vast majority of the infected are asymptomatic, the last two statistics, daily/cumulative infected, are, in reality, not directly observable by the policy maker. Fig. 3 further examines the corresponding economic measures, namely, hospitalization rates (proxied via the number of critically and severely daily infected individuals) and death rates.

**Table 1:** Cumulative percentages of infected across different compliance level and vaccination rate^†^

| Scenarios | Testing Policy | Random Vaccination |  | No Vaccination |  |
| --- | --- | --- | --- | --- | --- |
|  |  | %Population | SE | %Population | SE |
| Low Compliance Level<br>Low Vaccination Rate | Contact Tracing (CT) | 22.8311 | (0.5748) | 46.0067 | (0.1907) |
|  | CT + Random | 22.2713 | (0.4952) | 44.6116*** | (0.1952) |
|  | CT + Active Sample | 17.5112*** | (0.5343) | 41.3715*** | (0.2123) |
| Low Compliance Level<br>High Vaccination Rate | Contact Tracing (CT) | 11.6922 | (0.5338) |  |  |
|  | CT + Random | 10.3761* | (0.5569) |  |  |
|  | CT + Active Sample | 8.4913*** | (0.4547) |  |  |
| High Compliance Level<br>Low Vaccination Rate | Contact Tracing (CT) | 18.1235 | (0.5831) | 37.0027 | (0.3491) |
|  | CT + Random | 16.2956** | (0.4838) | 33.9982*** | (0.2088) |
|  | CT + Active Sample | 10.1984*** | (0.3891) | 27.4378*** | (0.3999) |
| High Compliance Level<br>High Vaccination Rate | Contact Tracing (CT) | 9.1383 | (0.5171) |  |  |
|  | CT + Random | 8.1184<sup>+</sup> | (0.4622) |  |  |
|  | CT + Active Sample | 5.9998*** | (0.3738) |  |  |

<sup>†</sup>%Population column reports the mean percentage of population infected under different compliance levels (10% and/or 50%) and different vaccination rates ( $\sim 0.25\%$ and/or $\sim 0.5\%$ per day), averaged over 50 simulation runs. The corresponding standard errors (SE) are reported in parentheses.
Significance levels are marked relatively to CT.
Significance level: <sup>+</sup> $p < 0.1$ , \* $p < 0.05$ , \*\* $p < 0.01$ , \*\*\* $p < 0.001$ .

**Table 2:**
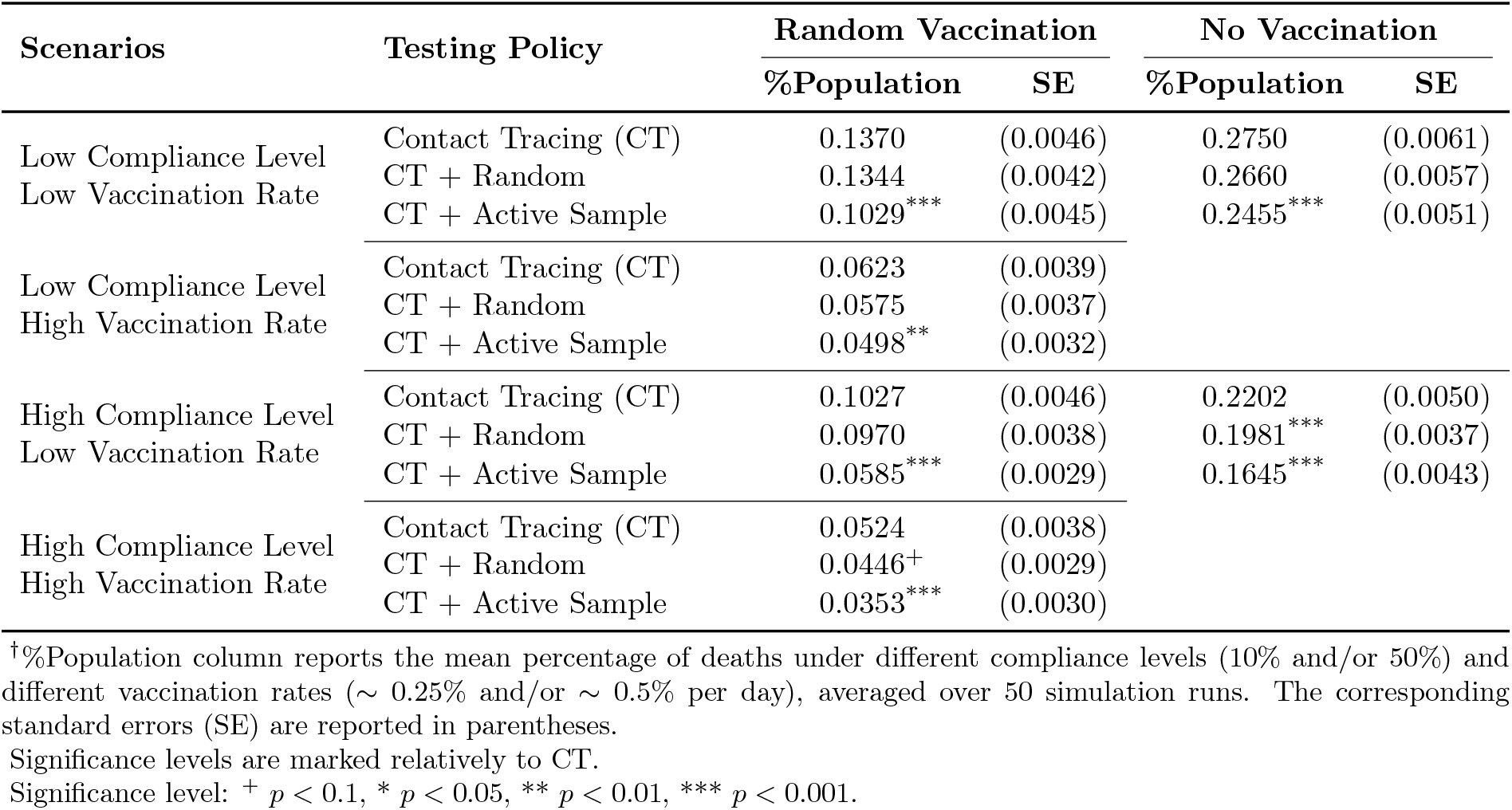
Cumulative percentages of deaths across different compliance level and vaccination rate^†^

| Scenarios | Testing Policy | Random Vaccination |  | No Vaccination |  |
| --- | --- | --- | --- | --- | --- |
|  |  | %Population | SE | %Population | SE |
| Low Compliance Level<br>Low Vaccination Rate | Contact Tracing (CT) | 0.1370 | (0.0046) | 0.2750 | (0.0061) |
|  | CT + Random | 0.1344 | (0.0042) | 0.2660 | (0.0057) |
|  | CT + Active Sample | 0.1029*** | (0.0045) | 0.2455*** | (0.0051) |
| Low Compliance Level<br>High Vaccination Rate | Contact Tracing (CT) | 0.0623 | (0.0039) |  |  |
|  | CT + Random | 0.0575 | (0.0037) |  |  |
|  | CT + Active Sample | 0.0498** | (0.0032) |  |  |
| High Compliance Level<br>Low Vaccination Rate | Contact Tracing (CT) | 0.1027 | (0.0046) | 0.2202 | (0.0050) |
|  | CT + Random | 0.0970 | (0.0038) | 0.1981*** | (0.0037) |
|  | CT + Active Sample | 0.0585*** | (0.0029) | 0.1645*** | (0.0043) |
| High Compliance Level<br>High Vaccination Rate | Contact Tracing (CT) | 0.0524 | (0.0038) |  |  |
|  | CT + Random | 0.0446<sup>+</sup> | (0.0029) |  |  |
|  | CT + Active Sample | 0.0353*** | (0.0030) |  |  |

<sup>†</sup>%Population column reports the mean percentage of deaths under different compliance levels (10% and/or 50%) and different vaccination rates ( $\sim 0.25\%$ and/or $\sim 0.5\%$ per day), averaged over 50 simulation runs. The corresponding standard errors (SE) are reported in parentheses.
Significance levels are marked relatively to CT.
Significance level: <sup>+</sup> $p < 0.1$ , \* $p < 0.05$ , \*\* $p < 0.01$ , \*\*\* $p < 0.001$ .

**Figure 2:**
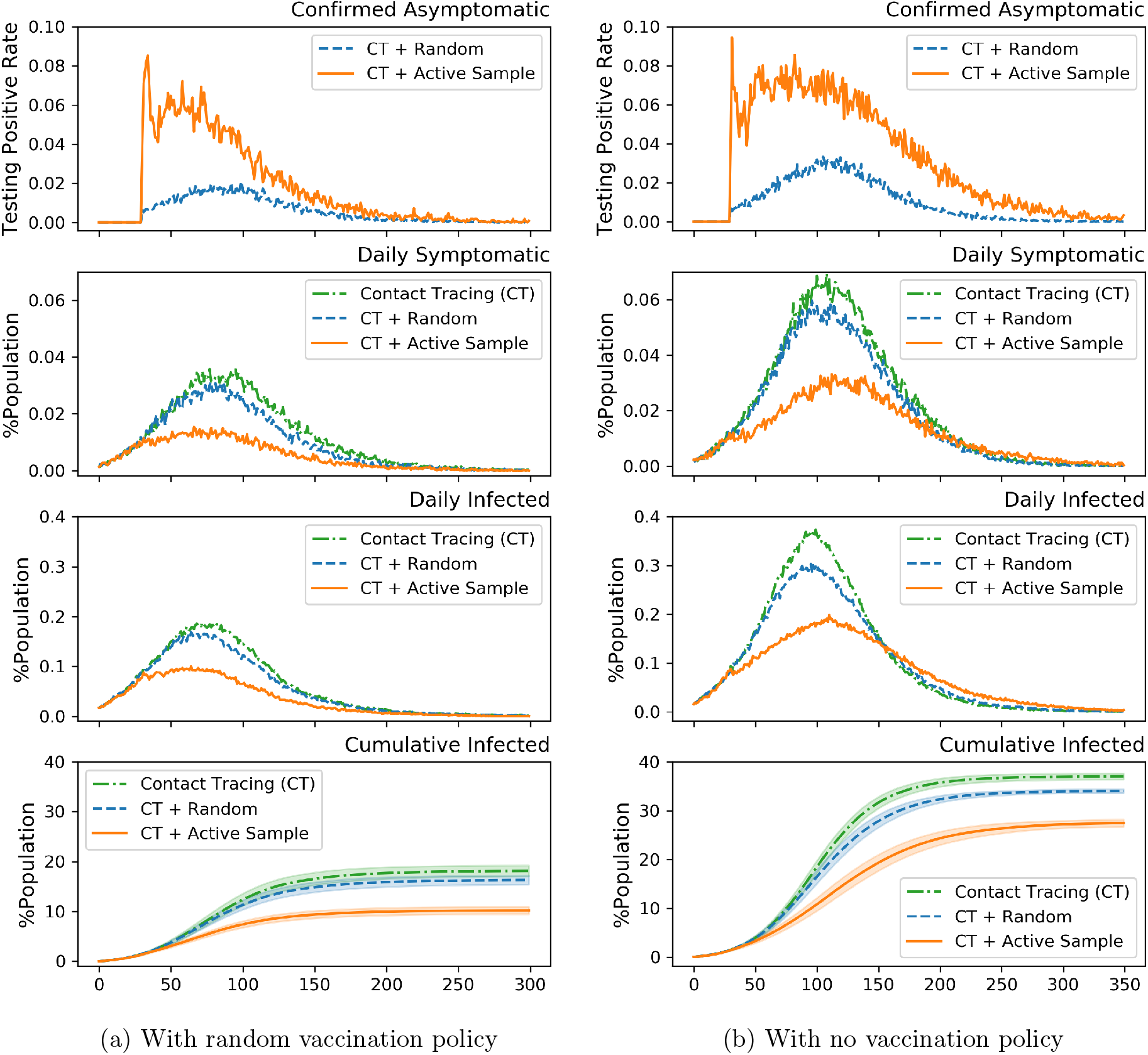
[Infections] Comparison between three testing policies, under high compliance level (*γ* = 50%) and low vaccination rate (*κ* = 25%). Error bands represent the 95% confidence interval.

**Figure 3:**
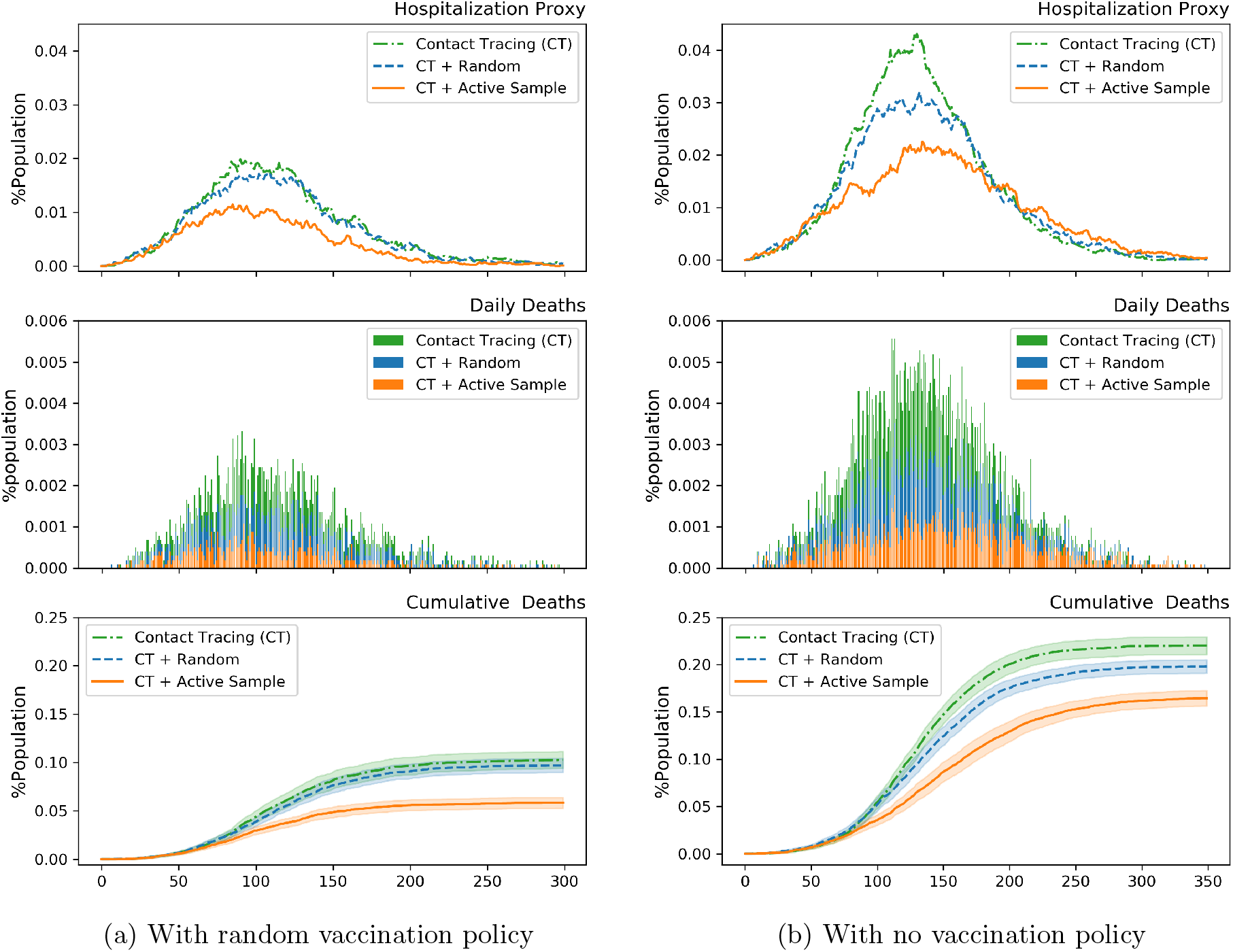
[Hospitalization and Deaths] Comparison between three testing policies, under high compliance (*γ* = 50%) and low vaccination rate (*κ* = 25%). Hospitalization is proxied based on how many are critically ill on each given day. Daily deaths are depicted in stacked bars. Error bands represent the 95% confidence interval.

Table 1 and Table 2 reports the full comparisons of three testing policies, i.e. contact tracing (CT), CT + Random, and CT + Active Sample, under four compliance/ vaccination scenarios, and without vaccination policy in place.

The main takeaways from these results are the following:

- Even with high vaccination rate, active sampling is still useful in reducing the extent of the disease. Specifically (from Tables 1 and 2), the death rate significantly drops by 20% to 30%; the percentage of infected drops by approximately 30%. These numbers are very significant. In the US population, these numbers map to approximately 10M fewer cases and 150K fewer deaths. With low vaccination rate, the effect of active sampling is even more notable. For instance, with low vaccination rate and high compliance, the percentage infected drops by ∼ 45%.
- Asymptomatic transmission is a significant aspect of Covid-19. Our results show that smart testing strategies generate 4x the test positivity rate as random testing. Catching the asymptomatic carriers early is a critical part of curbing the spread and decreasing infection/death rates; our active sampling method is effective at doing this, which is a key reason for its effectiveness.
- Load on hospitals at peak times, a crucial aspect of managing Covid-19, drops, according to Fig. 3, by 50% when implementing smart testing (with or without vaccinations).
- While the focus of this paper in not entirely on vaccination, the results emphasize the importance of vaccination. Even with a low vaccination rates, the death rate can drop by over 50% (see Table 2).
- Is random testing useful? Our results provide mixed evidence. It is useful at identifying asymptomatic individuals when the spread is large, but with reasonably large vaccination numbers don’t necessarily help in significantly lowering death rates. In pockets where compliance rates are high, the spread is increasing, but vaccination rates are low (perhaps due to availability), random testing is likely to be an effective complement to contact tracing alone.
- One major unknown is the extent of compliance in the population (research has shown that this varies from low single digits to as high as 100%) to self-quarantining requirements. The results show that testing policies are less effective when compliance is low. This is not surprising, since one of the main advantages of early identification of asymptomatic individuals is cutting the infection chain through quarantine policies. When the compliance is low, this advantage diminishes significantly. One possibility for policy, which we turn to next, is to consider compensation strategies that can increase compliance (Bodas and Peleg 2020).

## 5 Conclusion

This paper presented a smart-testing approach that proactively identifies individuals to test, concurrent with vaccination policies, to mitigate the spread of pandemics such as COVID-19. With increasing vaccination worldwide the focus on testing has unfortunately reduced. Our results clearly demonstrate the importance and urgency of adding smart-testing to current vaccination strategies for combating the pandemic.

Our method can immediately be used by policy makers to determine whom to test if the following information can be provided: (1) a list of all individuals in the population; (2) the set of known infected individuals in the population; (3) daily-contact network for individuals - ideally, this should be provided for all individuals that were tested positive, but the method can work even with partial information (daily contact network, for instance, can be computed easily from readily available mobile tracking data), and (4) if available, a partial mapping of individuals to locations that they can be associated with (e.g., household, workplaces, schools).

Our method can also be used by policy makers to examine carefully constructed scenarios in a world with smart-testing strategies. These scenarios may include different quarantine policies, intervention strategies or social behavior such as compliance or willingness to be vaccinated. The scenarios constructed can also experimentally then show the outcomes under smart-testing versus other testing strategies.

## Data Availability

This is a simulated study.

## Appendix

### A Agent-Based Model

In the real-world setting, as long as we have daily spatio-temporal data on individuals in a population, the algorithm presented in the previous section can be used to select individuals to test. However, and somewhat counter-intuitive, real-world data alone is insufficient to fully understand the impact of different sampling policies. The “real world” represents a single run of how reality is shaped, and does not offer the advantage of running experiments to test counterfactuals. For instance, would the sampling algorithm have worked as well under a different quarantine policy, or under various compliance scenarios? Agent-based models, on the other hand, are particularly effective to answer these types of more general questions.

In this paper we couple a disease progression model (Hethcote 2000) with Agent Based simulation Model (ABM) (Perez and Dragicevic 2009) of infectious spread under different settings. Disease progression is commonly modeled with the deterministic compartmental S/I/R (Hethcote 2000, Cooper et al. 2020), which divides the population into three compartments - susceptible to the disease (S), actively infected with the disease (I), and recovered (or dead) and no longer contagious (R)) - and defines transmission rates between the compartments. To account for additional aspects of the Covid-19 transmission, we use an extended S/I/R model that includes individuals that are exposed (E) to the disease (Silva et al. 2020, Prem et al. 2020), and asymptomatic (A) individuals (Manchein et al. 2020, Koo et al. 2020) - an S/E/A/I/R model.

We constructed a data driven stochastic Agent-Based Model (ABM) of the COVID-19 epidemic in New York city. Based on Hoertel et al. (2020), our ABM model includes four components: (1) synthetic population with demographic characteristics and spatial information that is generated to resemble the city of New York, (2) daily interaction network between individuals (agents) in the population, (3) disease dynamics that spreads via interactions, and progresses as an S/E/A/I/R model, (4) policy makers, as agents, who influence the environment based on the sampling strategies and quarantine policies in place.

In the reminder of this section we describe components (2) - (4) of the ABM. The population generating approach in component (1) is detailed in Appendix B. In Algorithm 1 we provide the pseudo code of our model.

### A.1 Interaction data structure

Our data is composed of a set of individual agents *A* = {*a*_1_, …, *a*_*n*_}, a set of geographic locations *L* = {*l*_1_,..*l*_*m*_}, and the propensity of the interactions between the agents and the locations: 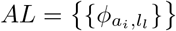 (how often an agent goes to the location).

#### Algorithm 1: Pseudo-code for the ABM

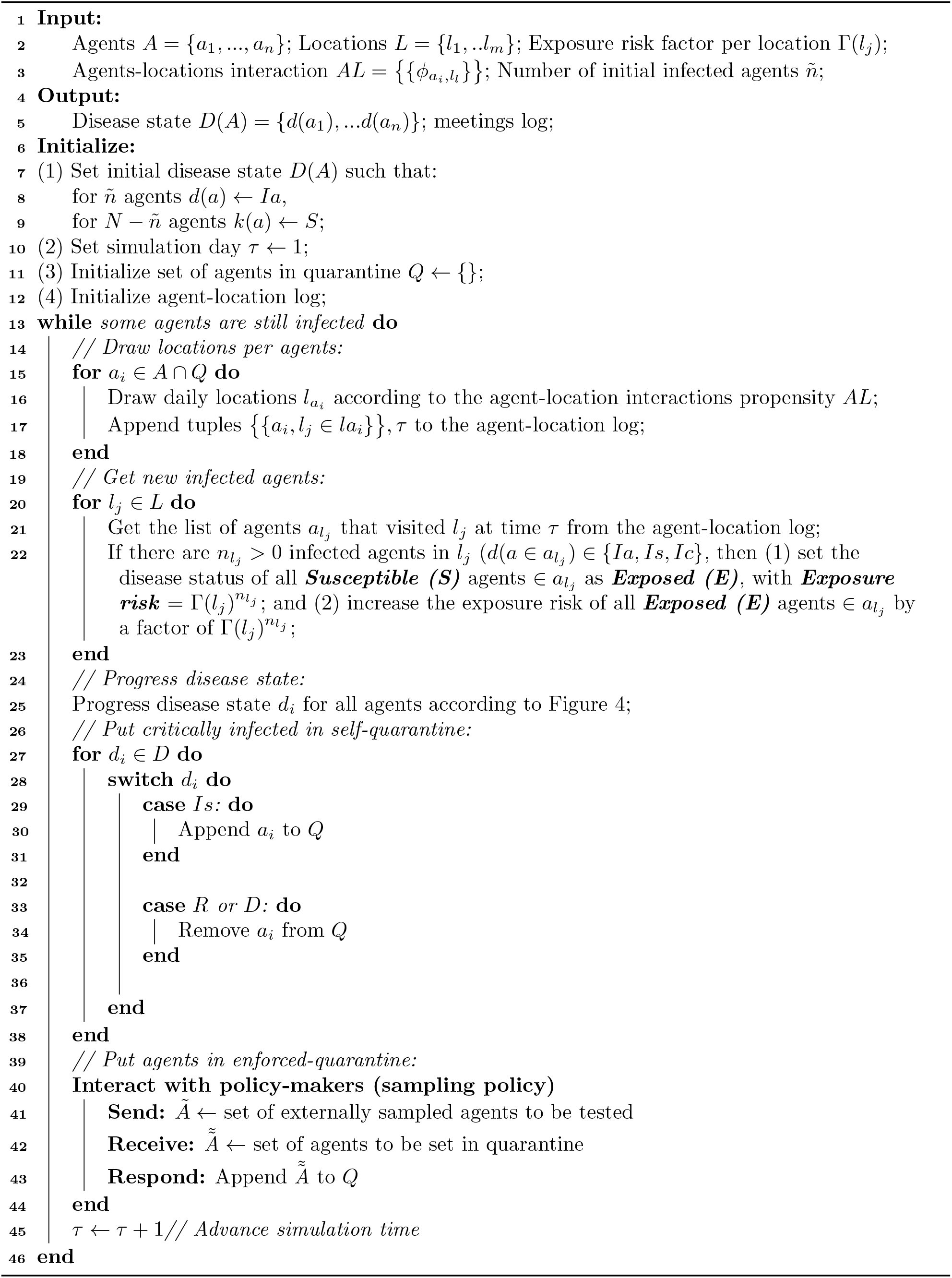

Agents in our data is characterized by age, gender, and a list of locations that the agents goes to, at a given probability. Locations can take different location types, and are spread across the neighborhoods of New York City. For this paper we set the following location types: household, workplace, school, station, and supermarket. Two agents may meet if they go to the same location at the same time. The meeting probability at a given location is a function of the location type (for example, agents that live at the same house will meet with a probability of *ϕ*_*household*_ = 1, while agents that go the the same supermarket, will meet at a much lower probability: *ϕ*_*supermarket*_ *<<* 1). In addition to the different location types, we add a special location type that we call a “mixing location”, in which agents can meet at random. Mixing locations in our model simulates meeting at public spaces, such at parks, shops, theater, etc.

Note that our sampling algorithm works as an agent within this framework and can be used with any ABM setting - all that is required is for the ABM to simulate contacts and movements of individuals in a population. The above are used primarily for exposition to simulate one specific city (New York). Agents in our model can potentially be in one of seven disease states, as described below. We formally define *D*(*A*) = {*d*(*a*_1_), …*d*(*a*_*n*_)} to be the set of disease states of all agents.

### A.2 Disease dynamic

The disease process used in this paper follows the deterministic *S/E/A/I/R* (Susceptible (*S*) – Exposed (*E*) - Asymptomatic (*A*) - Infectious (*I*) - Recovered (*R*)) compartmental model, with death (*D*) rate. We further split the infectious state into three (instead of two) disease stages: Infected asymptomatic (*Ia*), Infected symptomatic (*Is*), and Infected critical (*Ic*). Beyond being a more accurate description of COVID-19, this allows us to flexibly define different policies and different behaviours to infected individuals according to the state of their disease. Similar to *S/E/A/I/R*, we assume Recovered (*R*) is an absorbing state, implying that infected individuals can either die or become immune. Figure 4 presents the disease states and the transmission dynamics in this paper.

**Figure 4:**
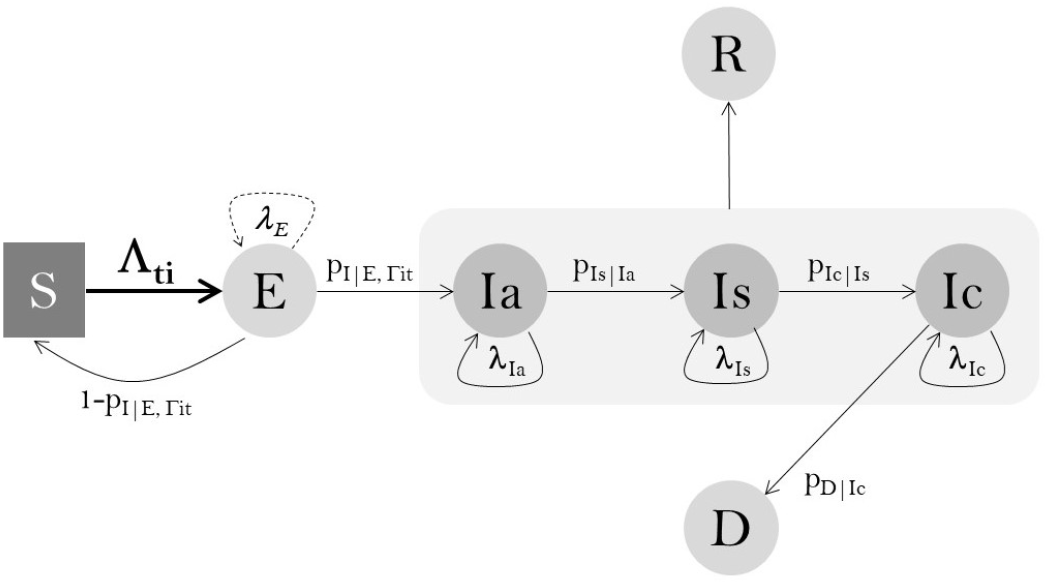
Transmission dynamics of the Corona virus

Transmission probabilities are assumed to be constant across individuals. This is due to the fact that the impact of patient’s covariates on the disease dynamics is still unknown. An exception is the transmission rate between states *S* and *E* – Λ_*it*_ – that determines the rate at which uninfected individuals are exposed to infected individuals, and depends on the individuals’ ego-network structure. Once exposed, they become infected with probability of 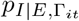. The argument Γ_*it*_ indicates the exposure risk factor, which depends on the number of infected agent that individual *i* met at time *t*, and the strength of her relationship with them. Specifically, following Madewell et al. (2020) we differentiate the probability of being infected infected from close connectors (a total of 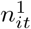), such as house mates or colleagues, from the probability of being infected from random contacts (a total of 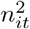), such as meetings at supermarkets, or metro stations: 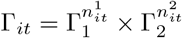

In infected individuals, the disease may escalate and become mild (*Is*), at a probability of *p*_*Is*|*Ia*_, then critical (*Ic*), at a probability of *p*_*Ic*|*Is*_. At any stage of the disease, infected patient can recover. Recovery takes on average *λ*_*Ia*_ for asymptomatic agents, *λ*_*Ia*_ + *λ*_*Is*_ for symptomatic agents, and *λ*_*Ia*_ + *λ*_*Is*_ for critically ill agents. The latter disease state can result in death, at a rate of *p*_*D*|*Ic*_.

In the results presented in the paper, we set *λ*_*Ia*_ = *λ*_*Is*_ = *λ*_*Ic*_ = 0.9, implying an average of 10 days stay at each infection state (following the average infection period reported by Lauer et al. (2020)). Due to the difficulties in differentiating incubation period from asymptomatic period, as reported by Roda et al. (2020), we set *λ*_*E*_ = 0, effectively merging states *E* and *Ia*. The transition probabilities used in our results are: Γ_1_ = 0.02, Γ_2_ = 0.004, *p*_*Ic*|*Is*_ = 0.006, and *p*_*D*|*Ic*_ = 0.05. The numbers were set according to the disease statistics, as reporter by the World Meter (World Meter 2020).

We note that the sampling algorithm presented in this paper only uses data observed as a result of the actual disease diffusion process, and does not depend specifically on the details of how this is implemented in the ABM. In that sense, our approach will work with any extensions of the model implemented in here.

### A.3 Policy makers as agents

At the end of each ABM simulation epoch we sample a set of agents to be tested for COVID-19. Once an agent is sampled, her disease state as well as (possibly incomplete) contact network are revealed to policy makers, whom can then decide which agents should be put into quarantine. Other than sampled agents, we assume that critically infected agents self quarantine themselves. The quarantine duration is set for 14 days, following the current global practice. While in quarantine, an agent’s disease dynamics continues, but her interactions with others stop.

## B Data Structure of ABM

In this section we describe how we generated the synthetic population for the ABM. The synthetic data was generated to describe the city of New York. Specifically, we focus on the Manhattan area, in which the population is large and dense.

### B.1 Individual characteristics

Agents in our model were characterized by age and gender, both follow the age and gender distributions in NYC (United States Census Bureau 2020), as depicted in Table 3. These characteristics were later used to define social interactions according to household structures in NYC, age groups, etc.

**Table 3:** Individual Characteristics

|  | Level | Percentage |
| --- | --- | --- |
| Age | <5 | 6.8% |
|  | 5-9 | 7.0% |
|  | 10-14 | 6.6% |
|  | 15-19 | 6.5% |
|  | 20-24 | 7.4% |
|  | 25-34 | 17.1% |
|  | 35-44 | 15.8% |
|  | 45-54 | 12.6% |
|  | 55-59 | 4.6% |
|  | 60-64 | 3.9% |
|  | 65-74 | 6.2% |
|  | 77-84 | 4.0% |
|  | >85 | 1.5% |
| Gender | Male | 47.38% |
|  | Female | 52.62% |

The number of agents varied in our simulation runs between (1) the entire Manhattan population, which is approximately 2M, (2) a large subset of the population: ∼ 100K, (3) a small subset of the population: ∼ 20k.

### B.2 Geographic locations

The spatial layout included ten groups of adjacent Neighborhood Tabulation Areas (NTA’s) in Manhattan (NYC Open Data 2020). The reason for grouping NTAs stemmed from the granularity of the data that is available for the number of business in NYC (NYC Comptroller 2020).

NTAs in our data were virtually positioned in our spatial layout according to its central longitude and latitude. Each NTA included multiple locations of five types: households, workplaces, schools, stations, and supermarkets, which we located uniformly around the center. The number of households and workplaces in each NTAs were taken from NYC Open Data (2020) and NYC Comptroller (2020), respectively. A total of 1700 schools (NYC Department of Education 2020), and 470 stations (NYC Transit 2020), were evenly divided across NTAs. The number of supermarkets where computed based on one supermarket per each 100000 inhabitants statistics (Hoertel et al. 2020). Other location types, such as public parks, shops and theathers, were modeled via “mixing location”, as we explained in Appendix A.

Table 4 provides the list of NTAs, their coordinates, and the location types and numbers per NTA.

**Table 4:** Summary of NTAs in Manhattan

| NTA | *long.* | *lat.* | *household* | *schools* | *stations* | *markets* | *workplaces* |
| --- | --- | --- | --- | --- | --- | --- | --- |
| Battery Park City,<br>Greenwich Village &<br>Soho | 40.86183 | 73.92363 | 61672 | 170 | 47 | 35 | 18626 |
| Central Harlem | 40.8221 | 73.92363 | 48680 | 170 | 47 | 28 | 1270 |
| Chelsea, Clinton &<br>Midtown Business District | 40.8089 | 73.9482 | 59821 | 170 | 47 | 34 | 32137 |
| Chinatown &<br>Lower East Side | 40.7957 | 73.9389 | 67708 | 170 | 47 | 39 | 8573 |
| East Harlem | 40.7736 | 73.9566 | 48174 | 170 | 47 | 28 | 1721 |
| Hamilton Heights,<br>Manhattanville &<br>West Harlem | 40.787 | 73.9754 | 51826 | 170 | 47 | 30 | 1962 |
| Murray Hill,<br>Gramercy &<br>Stuyvesant Town | 40.75507 | 73.9924 | 62339 | 170 | 47 | 36 | 22156 |
| Upper East Side | 40.7388 | 73.97933 | 91671 | 170 | 47 | 53 | 8896 |
| Upper West Side &<br>West Side | 40.7154 | 73.99065 | 80736 | 170 | 47 | 46 | 6406 |
| Washington Heights,<br>Inwood &<br>Marble Hill | 40.72283 | 74.00717 | 81367 | 170 | 47 | 47 | 3049 |

### B.3 Social Interactions

Agents in the simulation were assigned to locations in the virtual grid. The visit frequency to assigned location depends on the location type, as we describe below. Interactions between agents occurred via visits to the same location.

The agents assignment to locations, and the meeting probability at each location type is as follows.

#### Households

Agents were assigned to households randomly according to the family structure in NYC (United States Census Bureau 2020). Daily meeting probability within each household was equal to 1.

#### Schools

kids of age 6-18 were assigned to the closest school to their household. Within each schools, kids were clustered into classes according to their age group, and average class size of 26 (NYC Department of Education 2020). Meeting probability within class was assumed to be 1 and 0 otherwise.

#### Workplaces

Adults of age 20-60 were assigned to Workplaces according to the NYC employment rate (95.9%, (NYC Department of Labor 2021)). Assignment was not geo-dependent. Following Hoertel et al. (2020), 89% of the workplaces were considered small. In these places, workers were clustered into working groups of three colleagues on average. Large workplaces accounted for 11% of all workplaces, with workers grouped into groups of 11 workers on average. Meeting probability with colleagues was assumed to be 1, and 0 with other workers.

#### Supermarkets

We assumed working agents go to supermarkets that are closest to their household and workplace. Non-working adult agents were assumed to choose the supermarket that is closest to their household. Visits to a supermarket occurred on average twice a week (assumption). Meeting probability of two agents that are assigned to the same supermarket was computed such that each agents interacts with an average of 30 other agents in a visit.

#### Stations

Working agents were assumed to use the subway stations that are closest to their household and workplace. Meeting probability of two agents that go to the same station is computed such that each agents interacts with an average of 10 other agents in a ride.

Table 5 summarizes the social interactions.

**Table 5:**
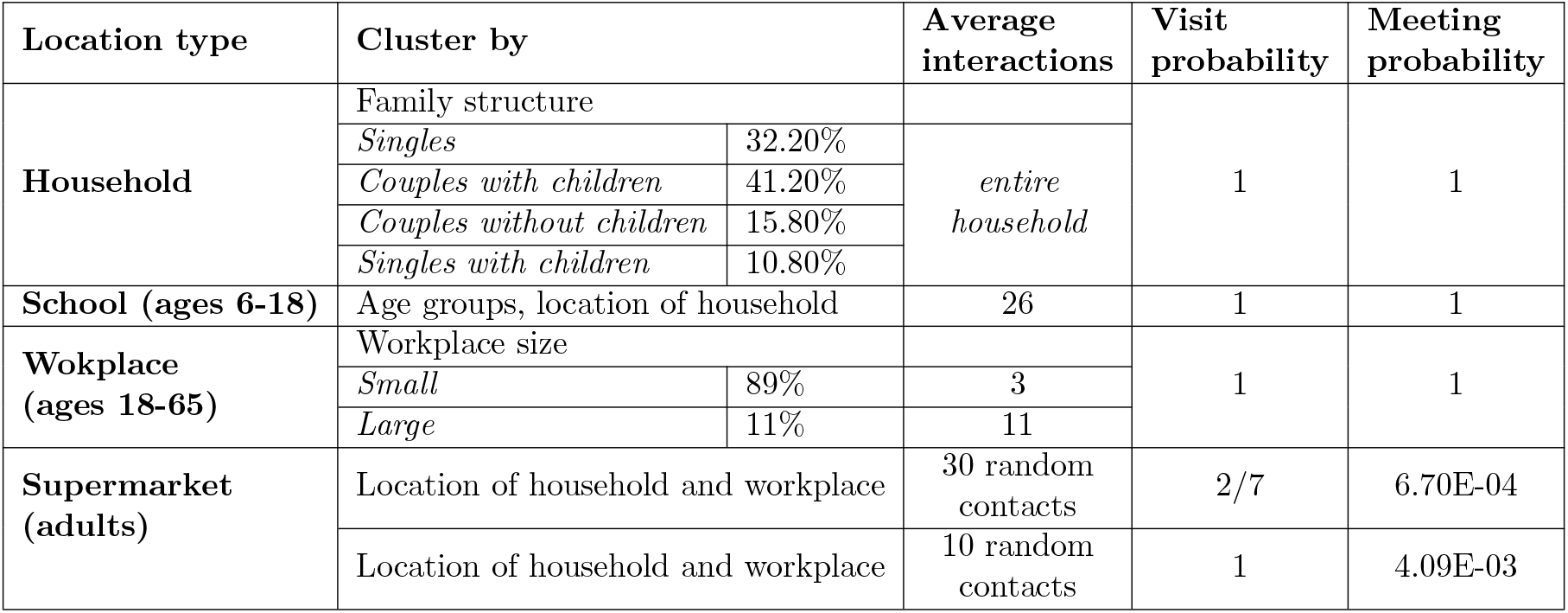
Summary of Social Interactions

## C DeepWalk

The below are the technical details of DeepWalk (Perozzi et al. 2014) - the use of language modeling approaches to learn latent node embedding in two steps: random walk and word embedding. The analogy made by DeepWalk is that nodes in a network can be thought of as words in a natural language.

### Random Walk on Graph

The first step of DeepWalk is to identify the context nodes for each node. Start at any given node, it identifies all its neighbors, randomly select one, and walk, with pre-defined path length, as illustrated in Fig. 5. Repeat until there are enough samples. By generating truncated random walks from the network, the context nodes of *v* ∈ *V* can be defined as the *k* neighboring nodes in each random walk sequence, which is a combination of nodes from *v*’s 1-hop, 2-hop,…, k-hop neighbors. The idea is that the techniques used to model natural language (where the symbol frequency follows a power law distribution) can be re-purposed for nodes appearing in short random walks whose frequencies also follow a power-law distribution.

**Figure 5:**
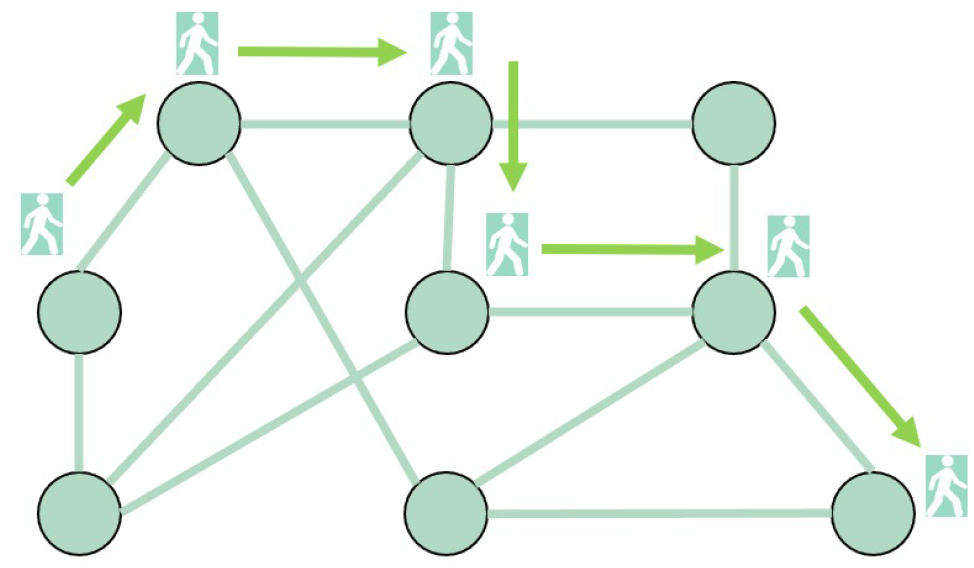
An example random walk of length 6 on a network.

### SkipGram

SkipGram algorithm is used to learn word embeddings which represent words in vector space so that if the word embeddings are close to one another, those words are semantically similar to one other (Mikolov et al. 2013). The key idea of SkipGram is to quantify the similarity between any two words by how frequently they share the same surrounding words. Skip-gram first identifies the context (neighboring) words for a given target word within a pre-defined window size. Then it predicts the context word by maximizing the conditional probability of observing the context words given the target word.

## D Supplement for Combinatorial *k*NN-UCB

We first begin with the mathematical proof of Theorem 2 that is used to modify the upper confidence bound for any 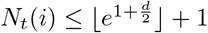.

### Theorem 3.

*For* ∀*c >* 0, *α >* 0, 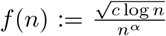 *is maximized at n*_0_ = *e*^1*/*2*α*^ *and is strictly decreasing for* ∀*n > n*_0_.

*Proof*. Based on the quotient rule, we take the derivative of 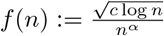, and yield:

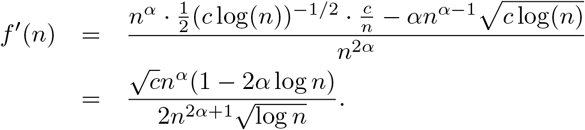

Since *c >* 0, *α >* 0, and *n >* 0, we can see that *f* (*n*) is maximized at *n*_0_ = *e*^1*/*2*α*^, strictly increasing when *n < n*_0_, and strictly decreasing after that.

We next present the pseudo-code for active sampling in Algorithm 2.

### Algorithm 2: Pseudo-code for Active Sampling (Combinatorial *k*NN-UCB)

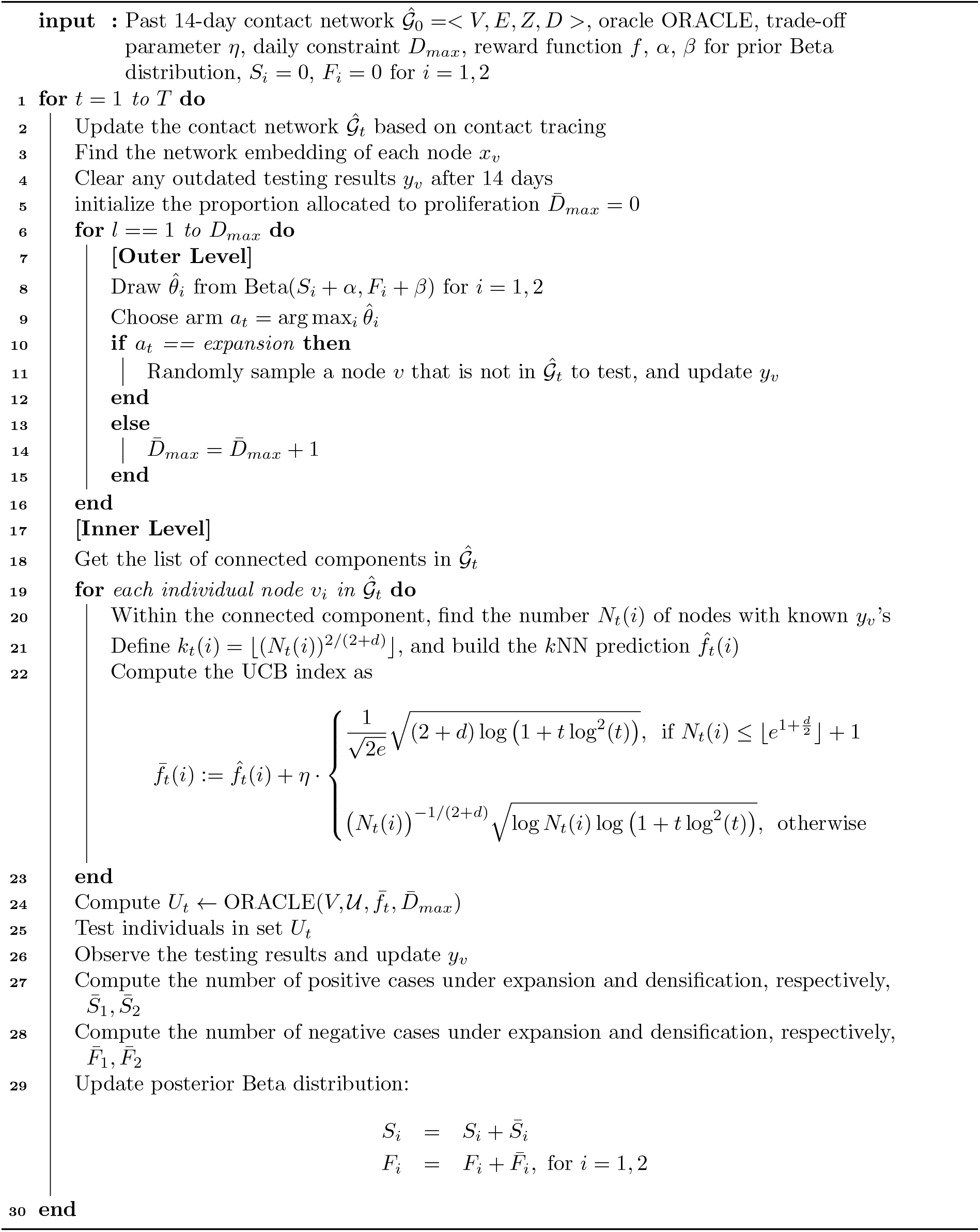

